# Prognostic Risk Refinement using Artificial Intelligence in HR+/HER2- Early Breast Cancer: Implications for CDK4/6 Eligibility Criteria

**DOI:** 10.64898/2026.01.23.26344621

**Authors:** Nicholas P McAndrew, Cynthia Ma, Andrew A. Davis, Elena Diana Chiru, Aditya Bardia, Jad M Abdelsattar, Joseph Cappadona, Ken Zeng, Krzysztof J. Geras, Jan Witowski, Cerise Tang

## Abstract

Patient selection and enrolment into phase III randomized clinical trials (RCTs) of adjuvant cyclin-dependent kinase 4 and 6 (CDK4/6) inhibitor therapies depend on accurate risk definition. However, standard clinicopathologic criteria incompletely capture recurrence risk, limiting their efficacy in treatment selection. To assess whether artificial intelligence (AI)-enhanced prognostication may enrich the clinical risk groups utilized in the adjuvant NATALEE trial, we evaluated Ataraxis Breast RISK (ATX), a multimodal AI test that integrates clinical data with morphological features from H&E-stained slides. ATX risk scores were generated for 2,228 patients with HR+/HER2- early breast cancer, of which 918 (41%) were classified as clinical high-risk and 1,310 (59%) were clinical low-risk. ATX was significantly associated with recurrence-free interval in both clinical risk groups and identified high-risk patients not captured by current clinical criteria, as well as individuals with limited benefit despite clinical high-risk classification. Consequently, integration of AI-enhanced risk assessment may improve selection of patients likely to benefit from adjuvant CDK4/6 inhibitors relative to current criteria.

## Introduction

For patients with early-stage hormone receptor-positive (HR+), human epidermal growth factor receptor 2-negative (HER2-) breast cancer, the standard of care has traditionally been surgery with or without radiotherapy and chemotherapy, followed by adjuvant endocrine therapy (ET) for 5 to 10 years with curative intent.^1^ Although adjuvant ET has significantly reduced recurrence risk in HR+/HER2- patients, 27-37% of patients with stage II disease and 46-57% of patients with stage III disease recur, sometimes up to 20 years or more post-diagnosis.^2,3^

Cyclin-dependent kinase 4 and 6 (CDK4/6) inhibitors, which impede cell-cycle progression through the cyclin D-CDK4/6-retinoblastoma pathway,^4–7^ have reshaped the treatment paradigm for HR+/HER2- breast cancer by being incorporated into adjuvant regimens. Two pivotal trials, NATALEE^8^ and monarchE^9–11^ have shown that ribociclib and abemaciclib, respectively, improve invasive disease-free survival among patients with stage II-III moderate- and high-risk HR+/HER2- early breast cancer.

The eligibility criteria for NATALEE included all node-positive and T3N0 patients, as well as T2N0 patients with one other high-risk factor, such as Ki-67 proliferation index ≥20%, grade III, or high genomic risk based on Oncotype DX,^12^ Prosigna PAM50,^13^ MammaPrint,^14^ or Endopredict.^3,15^ Patients meeting these criteria are considered to be at a higher risk of cancer recurrence and were identified as those most likely to benefit from the addition of an adjuvant CDK4/6 inhibitor. Recently published five year data demonstrated that patients in the ribociclib arm had significantly improved invasive disease-free survival as compared to the ET alone arm.^8^ However, eligibility criteria based on clinicopathologic definitions may offer a limited view of biological risk, which may lead to a subset of patients at a high biological risk to be misclassified as clinically low-risk. Moreover, other patients may meet enrollment criteria yet have biologically low-risk disease, so may experience limited or no benefit. These limitations underscore the need for improved risk stratification to guide adjuvant CDK4/6 inhibitor use.

To address the limitations of clinicopathologic risk stratification, we evaluated Ataraxis Breast RISK (ATX),^16^ an artificial intelligence (AI) test that integrates digital histopathology features from H&E-stained tumor slides with clinical features to generate individualized recurrence-risk estimates and refine the risk groups used in the NATALEE trial. We hypothesized that ATX could improve risk stratification over standard clinicopathologic variables, identifying refined patient subsets with varying levels of recurrence risk. Herein, we evaluate whether an AI-based model could stratify recurrence risk within current eligibility criteria.

## Results

### Risk stratification of HR+/HER2- early breast cancer patients across five datasets

To evaluate the performance of ATX, we assembled a multicenter cohort of breast cancer patients from five institutions. Of the 3,724 patients collected, 2,228 patients had confirmed early-stage HR+/HER2- tumors (**Figure 1**). Among these, 918 (41%) met the clinical risk criteria specified by the NATALEE trial and were classified as clinical high-risk, whereas 1,310 did not meet NATALEE criteria and were classified as clinical low-risk.

**Figure 1:**
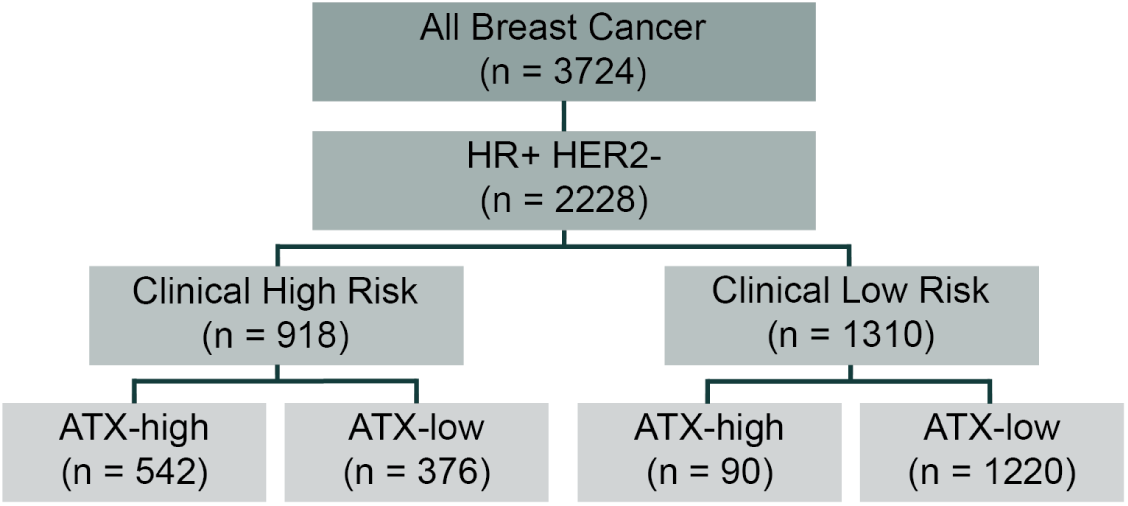
Study overview and cohort composition. Flowchart of patient selection starting with a total cohort of breast cancer patients across 5 institutional datasets (n = 3,724), narrowed by HR and HER2 status (n=2,228, 60%). Patients were then stratified by clinical risk based on NATALEE criteria, and ATX score. HR = hormone receptor, HER2 = human epidermal growth factor receptor 2.

ATX generated a 5-year risk of recurrence score for each patient (**Supplemental Figure 1**) and stratified patients into ATX-high and ATX-low groups. Within the clinical high-risk patients, 542 (59%) were concordantly classified as ATX-high while 376 (41%) were discordantly classified as ATX-low. Conversely, among clinical low-risk patients, 1,220 (93%) were concordantly classified as ATX-low, and 90 (7%) were discordantly classified as ATX-high.

The median follow-up was 5.08 years and 174 recurrence events were observed across all time points (event rate, 7.81%). Clinical high-risk and low-risk patients were comparable in age at diagnosis, menopausal status, and similar proportions of patients received adjuvant ET (**Table 1**). However, clinical high-risk patients had a higher average grade, tumor stage, and nodal stage. Clinical high-risk patients were more likely to have received adjuvant chemotherapy (45.8%) compared to clinical low-risk patients (12.8%). The median ATX score in clinical high-risk patients was 0.11 (IQR = 0.08-0.18) vs 0.05 (IQR = 0.04-0.06) in clinical low-risk patients.

**Table 1:** Patient Characteristics by clinical risk group.

| Characteristic | Category | Clinical High-Risk | Clinical Low-Risk |
| --- | --- | --- | --- |
| **Menopausal Status** | Peri-menopausal | 20 (4.5%) | 21 (7.4%) |
|  | Post-menopausal | 301 (68.3%) | 191 (67.7%) |
|  | Pre-menopausal | 120 (27.2%) | 70 (24.8%) |
| **Grade** | 1.0 | 58 (7.9%) | 219 (23.8%) |
|  | 2.0 | 378 (51.5%) | 568 (61.6%) |
|  | 3.0 | 298 (40.6%) | 135 (14.6%) |
| **T Stage** | T0 | 1 (0.1%) | 0 (0.0%) |
|  | T1 | 263 (28.6%) | 1177 (91.0%) |
|  | T2 | 483 (52.6%) | 106 (8.2%) |
|  | T3 | 147 (16.0%) | 1 (0.1%) |
|  | T4 | 24 (2.6%) | 1 (0.1%) |
|  | TX | 0 (0.0%) | 9 (0.7%) |
| **N Stage** | N0 | 180 (19.6%) | 1269 (98.2%) |
|  | N1 | 590 (64.3%) | 0 (0.0%) |
|  | N2 | 97 (10.6%) | 0 (0.0%) |
|  | N3 | 51 (5.6%) | 0 (0.0%) |
|  | NX | 0 (0.0%) | 23 (1.8%) |
| **Years at DX** |  | 57.7 (27-90) | 60.0 (29-90) |
| **Adjuvant Chemotherapy** | No | 279 (54.2%) | 748 (87.2%) |
|  | Yes | 236 (45.8%) | 110 (12.8%) |
| **Adjuvant Endocrine Therapy** | No | 40 (7.7%) | 78 (9.1%) |
|  | Yes | 479 (92.3%) | 778 (90.9%) |
| **ATX Risk Score** |  | 0.11 (0.08-0.18) | 0.05 (0.04-0.06) |
| **ATX Risk Category** | Low | 376 (41.0%) | 1220 (93.1%) |
|  | High | 542 (59.0%) | 90 (6.9%) |

Data are presented as n (%), except for ATX risk score, which is shown as median (IQR).

### ATX risk scores meaningfully stratify patients by recurrence risk

To determine the efficacy of ATX at stratifying patients by recurrence risk, recurrence-free interval (RFI) was used as the primary endpoint. Across all 2,228 HR+/HER2- patients, when stratified by risk score, there was a significant difference in the probability of meeting the RFI endpoint between the two groups (HR = 4.66, p_cox_ < 0.001), where ATX-high patients had a 5-year probability of 0.83 (95% CI: 0.77-0.86) and ATX-low patients’ probability was 0.95 (95% CI: 0.94-0.97) (**Figure 2A**).

**Figure 2:**
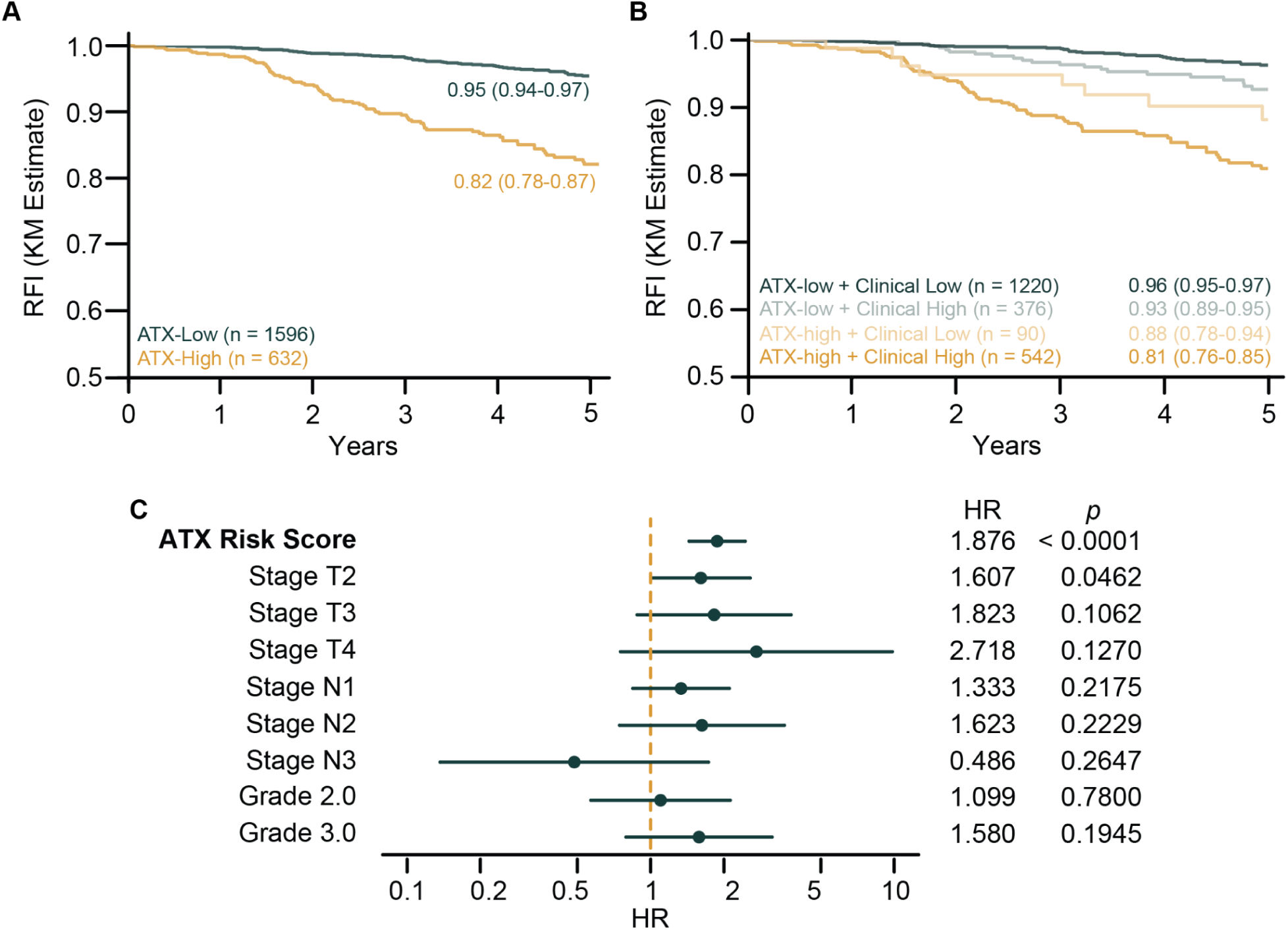
ATX RISK scores predict prognosis in HR+/HER2- patients. A) Kaplan-Meier plot of Recurrence-free interval in the full 2,228 patient cohort, stratified by ATX risk score (cut-off = 0.1). Recurrence-free interval at 5-years and corresponding 95% confidence interval are shown. B) Kaplan-Meier plot of recurrence-free interval in the full 2,228 patient cohort, with patients separated by ATX risk score (cut-off = 0.1) and clinical risk category. C) Multivariate Cox regression demonstrating that ATX risk score (by 0.1 increments) is associated with recurrence-free interval independent of other clinical factors.

Further stratifying patients by the combination of both ATX risk and clinical risk, we found that the four groups exhibited significantly different clinical outcomes (p_lrt_ < 0.001) (**Figure 2B**). Patients in the ATX-high + clinical low-risk group (n = 90, 4%) had lower 5-year probability of meeting the RFI endpoint (0.88, 95% CI: 0.78-0.94) than patients in the ATX-low + clinical high-risk group (n = 376, 17%; 0.93, 95% CI: 0.89-0.95).

Notably, the 5-year probability of remaining recurrence-free decreased progressively across the four risk strata from ATX-low + clinical low-risk (0.96, 95% CI: 0.95-0.97) to ATX-low + clinical high-risk (0.93, 95% CI: 0.89-0.95) to ATX-high + clinical low-risk (0.88, 95% CI: 0.78-0.94) to ATX-high + clinical high-risk (0.81, 95% CI: 0.76-0.85). We hypothesized that ATX risk score captured prognostic information beyond clinical features. To test this hypothesis, we fitted a multivariate Cox proportional hazards model including T stage, N stage, and grade as covariates stratified by dataset (**Figure 2C**). Across the full cohort of 2,228 HR+/HER2- patients, ATX score (HR = 1.88, 95% CI = 1.44-2.45, p < 0.001) and stage T2 (relative to stage T1, HR = 1.61, 95% CI = 1.01-2.56, p = 0.046) remained the only variables significantly associated with recurrence-free interval.

### Prognostic performance of ATX is consistent across cohorts

To assess the generalizability of ATX, we evaluated its prognostic performance across five independent cohorts. Hazard ratios and Harrell’s concordance-indices (C-index)^17–19^ were computed within each cohort and subgroup metrics were pooled using a random effects model. Among clinical high-risk patients, four out of five datasets showed a statistically significant association between ATX score and recurrence-free interval, and the pooled results were also statistically significant (HR = 1.92, p = 0.001) (**Figure 3A**). Similarly, the C-index confirmed strong discriminatory performance (C-index = 0.71, 95% CI = 0.63-0.77) (**Figure 3B**). In clinical low-risk patients, two of five datasets demonstrated statistically significant associations with ATX score, with the pooled result also significant (HR = 2.57, p = 0.016) (**Figure 3C**). The C-index was also strongly discriminatory (C-index = 0.70, 95% CI = 0.59-0.79) (**Figure 3D**).

**Figure 3:**
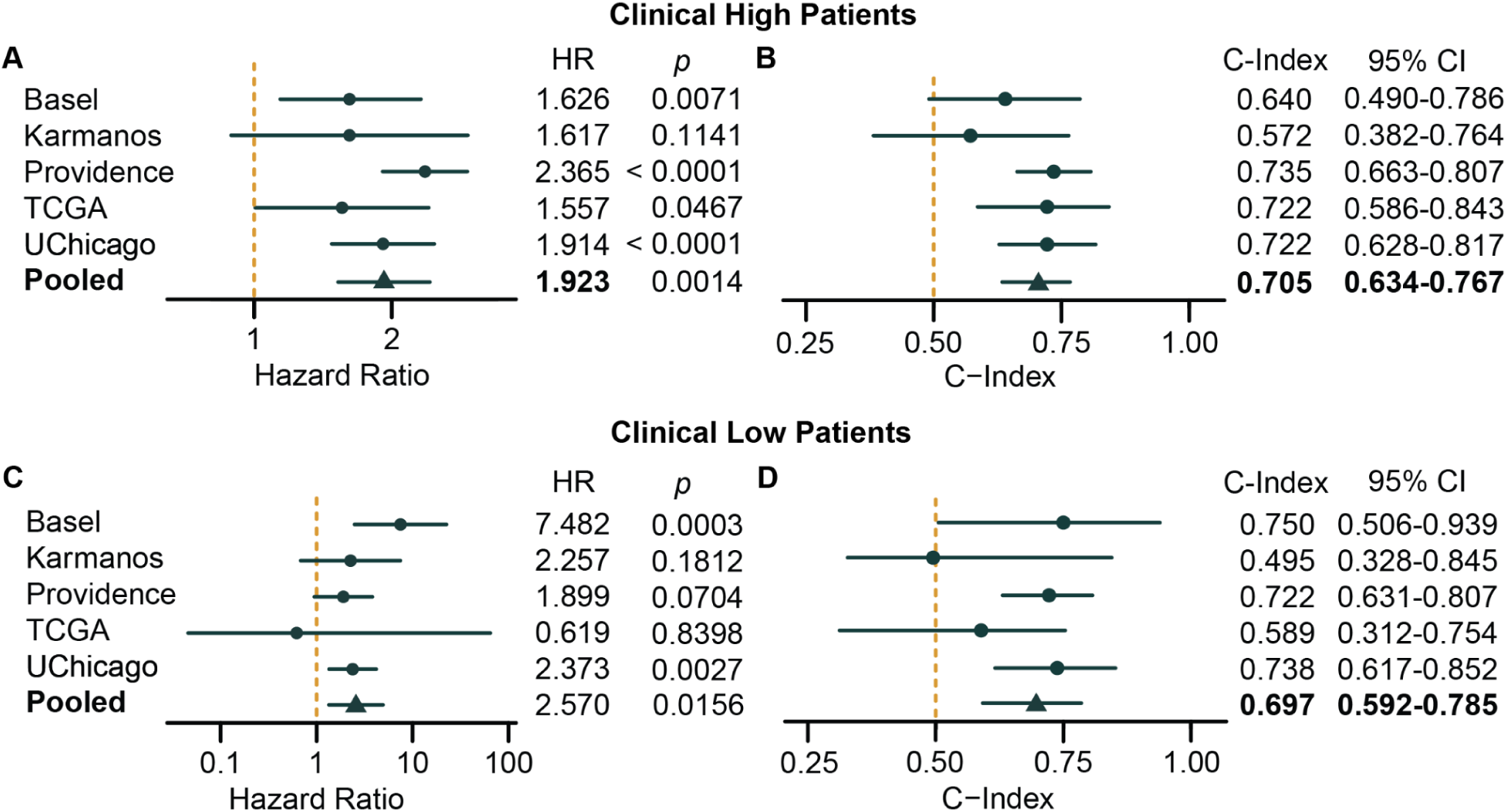
ATX performance is consistent across institutional cohorts and clinical risk groups. A) Prognostic ability of ATX in univariate models of clinical high-risk patients. Hazard ratio is calculated for each 0.1 increase in score. Results across datasets are pooled using a random effect model. B) Prognostic ability of ATX in clinical high-risk patients measured by c-index. Results across datasets are pooled using a random effect model and 95% confidence intervals are calculated using 1000 bootstrap iterations. C) Prognostic ability of ATX in univariate models of clinical low-risk patients. Hazard ratio is calculated for each 0.1 increase in score. Results across datasets are pooled using a random effect model. D) Prognostic ability of ATX in clinical low-risk patients measured by Harrell’s concordance index. Results across datasets are pooled using a random effect model and 95% confidence intervals are calculated using 1000 bootstrap iterations.

### ATX enhances risk stratification within clinical risk categories

We next sought to determine whether ATX meaningfully stratifies recurrence risk within clinically defined risk categories. Among clinical high-risk patients, those classified as ATX-high (mean KM-estimated 5-year RFI = 0.81, 95% CI = 0.76-0.85) had significantly shorter recurrence-free interval (HR = 3.55, p_cox_ < 0.001) than ATX-low patients (mean KM-estimated 5-year RFI = 0.93, 95% CI = 0.89-0.95) (**Figure 4A**). In a multivariate Cox proportional hazards adjusting for T stage, N stage, and grade stratified by dataset, ATX risk score remained the only variable significantly associated with RFI (HR = 1.90, p < 0.001) (**Figure 4B**) in clinical high-risk patients.

**Figure 4:**
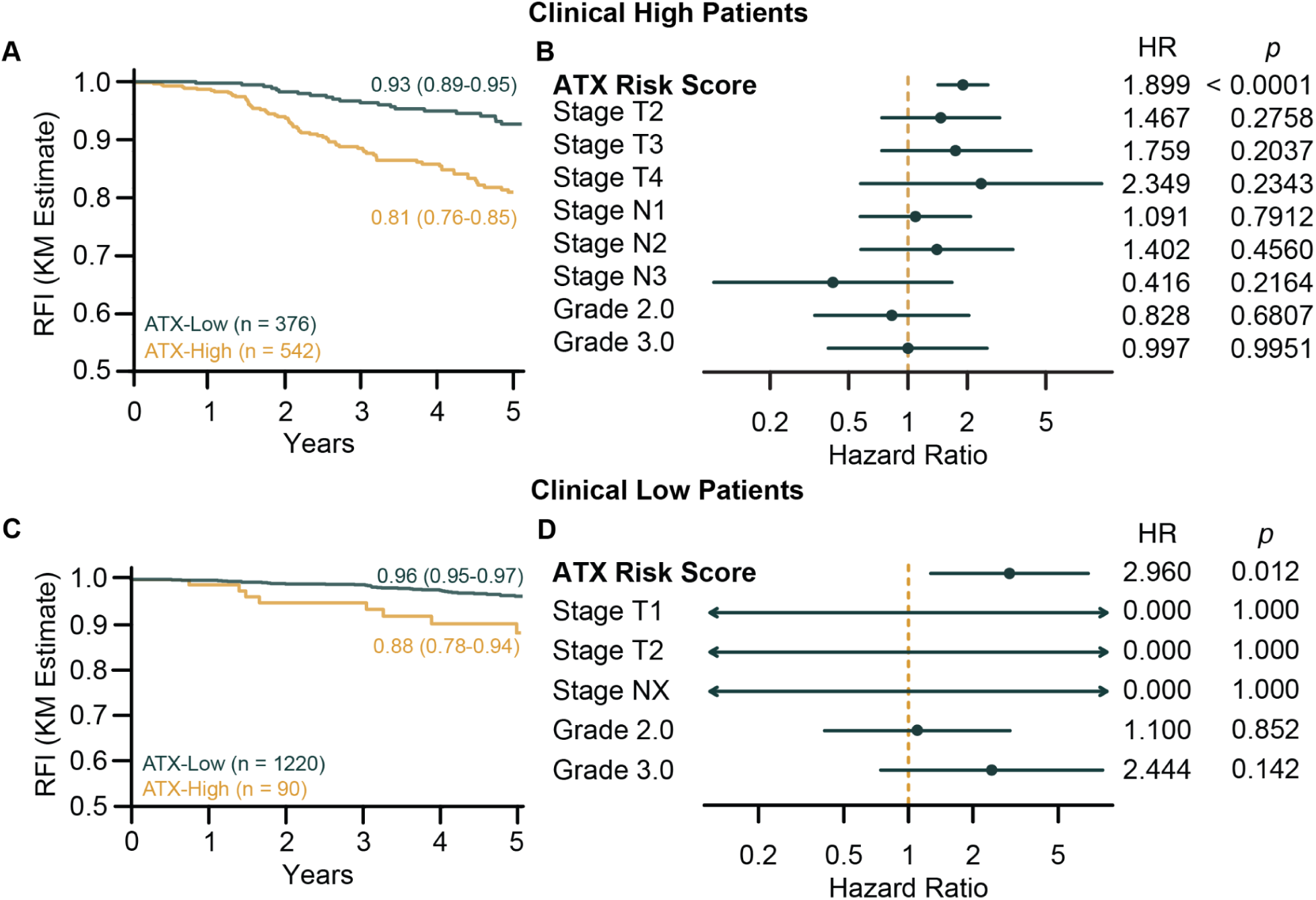
ATX RISK is prognostic within clinical risk categories. A) Kaplan-Meier plot of recurrence-free interval in clinical high-risk patients, stratified by ATX risk score (cut-off = 0.1). Recurrence-free interval at 5-years and corresponding 95% confidence interval are shown. B) Multivariate Cox regression of High-Risk patients demonstrating that ATX (by 0.1 increments) is associated with recurrence-free interval independent of T stage, N stage, and Grade. C) Kaplan-Meier plot of recurrence-free interval in clinical low-risk patients, stratified by ATX risk score (cut-off = 0.1). D) Multivariate Cox regression demonstrating that ATX risk score (by 0.1 increments) is associated with recurrence-free interval independent of other clinical factors in clinical low-risk patients.

Within clinical low-risk patients, ATX-high individuals (mean KM-estimated 5-year RFI = 0.88, 95% CI = 0.78-0.94) had significantly lower RFI (HR = 2.93, p_cox_ = 0.003) than ATX-low patients (mean KM-estimated 5-year RFI = 0.96, 95% CI = 0.95-0.97) (**Figure 4C**). Consistent with findings in the clinical high-risk cohort, the stratified multivariate Cox model also had ATX risk score as the sole independent predictor of recurrence-free interval (HR = 2.60, p = 0.012) (**Figure 4D**).

Further, within the clinical low-risk group, patients that are stage T2N0 disease are considered borderline eligible for ribociclib but require an additional risk factor to qualify. We aimed to determine if ATX could stratify T2N0 patients. ATX score-stratified patients demonstrated a visible separation in KM-estimated RFI between ATX-high and ATX-low groups but this analysis did not reach statistical significance (HR = 1.45, p_cox_ = 0.41) (**Supplemental Figure 2**).

### Prognostic utility of ATX is higher than clinical scores

To assess the potential clinical utility of ATX, we performed a time-dependent decision curve analysis (DCA) at five years, accounting for censoring using the Kaplan-Meier estimator. Decision curve analyses are depicted in **Figure 5A**. At lower recurrence risk thresholds for recommending treatment, the combined ATX score + clinical risk model conferred the highest net benefit, however as the threshold increased, ATX score alone outperformed both the combined model and clinical risk alone. Specifically, at an aggressive threshold of 0.05 (treating if chance of 5-year overall recurrence is > 5%), the net benefit of ATX score + clinical risk (0.041) was marginally higher than clinical risk (0.038) or ATX score alone (0.039) (**Figure 5B**). At 0.10, ATX score alone achieved the highest net benefit (0.025) compared to clinical risk (0.017) and ATX + clinical risk (0.018). This net benefit difference means that using the ATX score at the 0.10 threshold instead of NATALEE clinical criteria might avoid approximately 67 unnecessary treatments per 1,000 patients without compromising the number of appropriately treated recurrences. At a threshold of 0.15, ATX was the only strategy with positive net benefit (0.010), outperforming all other approaches and potentially avoiding approximately 55 unnecessary treatments.

**Figure 5:**
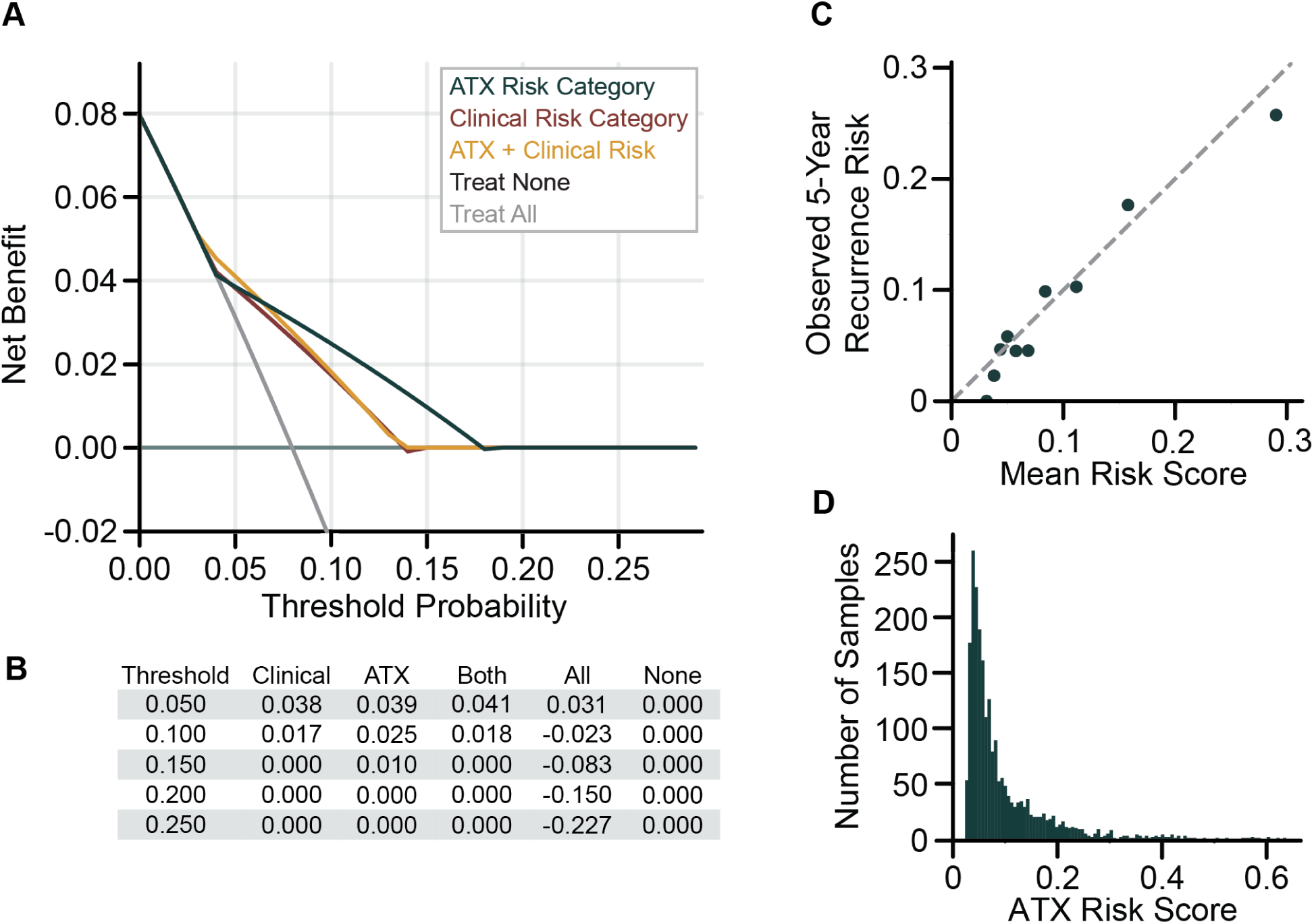
Clinical utility of ATX is greater than conventional clinicopathologic metrics. A) Decision curve analysis of the full 2,228 patient cohort comparing performance of ATX, clinical risk, and ATX combined with clinical risk. B) Table of net benefit values at specific thresholds for each model compared with treating all and treating no patients. C) Calibration analysis comparing observed 5-year event rate and mean risk score. D) Histogram of ATX risk scores across all 2,228 patients.

To ensure that ATX score predicted recurrence probabilities align with the actual observed outcomes in our patient cohort, we performed a time-dependent calibration analysis. We compared predicted 5-year recurrence probabilities based on ATX scores with observed event rates estimated using the Kaplan-Meier method within each risk decile (**Figure 5C**). Across all 2,228 HR+/HER2- breast cancer patients, ATX demonstrated close agreement between predicted and observed 5-year recurrence probabilities, with a Brier score of 0.067, indicating good overall calibration. Calibration was most accurate within risk scores below 0.3, which encompasses the vast majority of patients (97% of the study population) (**Figure 5D**). Together, these findings confirm that ATX provides well-calibrated and clinically interpretable risk estimates, reinforcing its utility in guiding adjuvant CDK4/6 inhibitor treatment decisions.

## Discussion

The NATALEE^3^ and monarchE^9–11^ trials established that CDK4/6 inhibitors improve disease-free survival among patients with stage II-III moderate- or high-risk HR+/HER2- early breast cancer. However, the true clinical impact of these trial findings has been questioned given the modest, albeit increasing with longer follow-up, absolute benefit and high rates of censoring in both trials.^20,21^ Moreover, current trial-based eligibility criteria rely on clinicopathologic features and select genomic assays that incompletely capture the biological heterogeneity of cancer. Consequently, some patients with high biological risk may be misclassified as low-risk and miss potential benefit from CDK 4/6 inhibitors, while others meeting clinical criteria may derive less benefit than expected. This imbalance underscores the need for refined, biologically informed approaches to risk refinement and treatment selection.

In a multicenter analysis, we demonstrated that ATX, a multimodal AI test integrating histology images with clinical data, provides independent, generalizable, and clinically relevant prognostic information in early HR+/HER2- breast cancer patients. ATX risk score was significantly associated with recurrence-free interval across five datasets and remained an independent predictor of recurrence after adjusting for T stage, N stage, and grade, underscoring that ATX score captures prognostic information beyond conventional clinical factors.

The enhanced risk stratification observed with ATX likely derives from its use of high-dimensional pathology embeddings extracted from H&E-stained slides. These representations capture spatial features such as tumor architecture, stromal composition, and local microenvironmental context, that are unavailable from genetic assays such as Oncotype DX. Importantly, a single bulk gene expression sample does not preserve spatial information and consequently is unable to capture the intratumoral heterogeneity of cellular activity or microenvironmental organization. By integrating morphological features with known clinical attributes, ATX bridges biological and clinical signals in its risk assessment offering a refined representation of tumor heterogeneity and behavior.

ATX stratified patients by recurrence risk within both clinical high and low-risk populations, supporting its value as a biologically informed prognostic test. Clinically, these findings suggest that ATX may identify high-risk CDK4/6 inhibitor-eligible patients who are not captured by existing clinical criteria, as well as patients who meet clinical high-risk criteria yet exhibit biologically low-risk disease and may potentially derive less than expected benefit. Together, these results establish that ATX provides robust and generalizable prognostic stratification across heterogeneous patient populations, suggesting its potential to refine adjuvant CDK4/6 inhibitor selection.

To evaluate potential clinical utility, we performed a time-dependent decision curve analysis comparing ATX score, clinical risk, and their combination. Decision curves quantify the net benefit of a prognostic model by balancing the value of correctly identifying patients who will recur against the potential harm of unnecessary treatment. Lower decision thresholds reflect a willingness to treat broadly to avoid missed recurrences, whereas higher thresholds prioritize specificity and reduction of overtreatment. At clinically relevant thresholds of 0.10 and 0.15, which reflect the recurrence risk observed in the NATALEE trial population (approximately 14% at 5 years in the control arm and 10% with ribociclib)^8^ and is consistent with the Kaplan-Meier-estimated 5-year cumulative recurrence risk in our dataset (8%), ATX demonstrated higher net benefit than clinical risk alone, indicating that ATX may offer greater clinical utility in guiding adjuvant CDK4/6 inhibitor decisions. Together, the decision curve analysis suggests that ATX scores may provide higher clinical usefulness than conventional clinicopathologic scoring at a moderate recurrence risk threshold.

Several limitations must be considered when interpreting the conclusions of our study. First, as this study utilizes real-world observational cohorts, cause-specific mortality was unavailable for all patients, hence the RFI used is an approximation rather than a strict implementation of the STEEP-defined endpoint^22^. Additionally, while the multicenter design enhances the generalizability of ATX, all cohorts were collected retrospectively. Further, ATX is a prognostic test, not a predictive test and whether ATX can identify patients who derive differential benefit from adjuvant CDK4/6 inhibitor treatment remains to be determined. Prospective validation in randomized clinical trials is necessary to confirm ATX’s predictive value and clinical utility. Moreover, information on adjuvant treatment was unavailable for a subset of patients in the study cohort. Finally, the median follow-up of our cohort was only 5 years so the full capability of ATX to capture recurrence at later timepoints may be attenuated.

In summary, this study suggests Ataraxis Breast RISK is a generalizable AI test for recurrence risk assessment in patients with HR+/HER2- early-stage breast cancer. By integrating clinical and histological features, ATX surpasses conventional clinical criteria in prognostic precision with strong calibration and clinical utility. In a time where some argue for expanded access to CDK4/6 inhibitors and others question their benefit and raise concerns over toxicity, our data suggests a complementary path where AI-based risk stratification may focus intensified therapy on truly biologically high-risk individuals while avoiding overtreatment in others. Therefore, our findings suggest ATX may potentially refine the risk groups eligible for adjuvant CDK4/6 inhibitors and advance biologically informed precision oncology.

## Methods

### Study design and population

A multicenter cohort of breast cancer patients from five institutions with available H&E slides and clinical information was assembled, totaling 3,724 patients with non-metastatic invasive breast cancer. After excluding 1,496 patients who were not HR+/HER2-, the final cohort included 2,228 patients. Patients were classified into clinical risk categories according to NATALEE criteria.^3^ Specifically, patients were considered clinical high-risk if they met any of the following:

1. Node positive disease (N1-N3) irrespective of tumor size (T0-T4).
2. T3N0 disease
3. T2N0 disease with an additional high-risk feature, defined as either grade 3 or Oncotype DX > 25.

Patients not meeting any of these criteria were classified as clinical low-risk. The primary endpoint in this study was an approximation of RFI, defined as duration from diagnosis to first recurrence of breast cancer, excluding death due to incomplete information on cause of death.

### Development of Ataraxis Breast RISK

The Ataraxis Breast RISK (ATX) test is multi-modal where its input consists of a digitized H&E slide and clinical variables (age, T and N stage, ER status, PR status, HER2 status, and histological subtype). First, morphological features from the slide are extracted using Kestrel, a vision transformer-based model trained on 400 million pathology images.^23^ Then, the features extracted by Kestrel and clinical variables are integrated and the probability of recurrence at 5 years is predicted. ATX was trained as a time-to-event model on over 4,500 samples. A higher ATX score suggests higher probability of experiencing cancer-related events.^16^ ATX is a class C1 prognostic biomarker based on 2025 European Society of Medical Oncology (ESMO) biomarker classification criteria.^24^ No data used for training ATX was included in this validation study.

### Time-dependent decision curve analysis

Time-dependent decision curve analysis (DCA)^25,26^ was performed using the dcurves package^27^ to compare 5 models at identifying patients with high risk of recurrence: 1) treat no patients, 2) treat all patients, 3) treated based on clinical risk criteria, 4) treat based on ATX criteria, and 5) treat based on a combined strategy (either clinical high-risk or ATX high). Univariate Cox proportional hazards models were fit to model time to recurrence separately for each model. The Cox proportional hazards models were then used to estimate the risk of recurrence for each patient and patients were considered test-positive at a given threshold probability if their risk of recurrence was greater than the threshold. True positive (TP) and false positive (FP) rates among test-positive patients were estimated using the Kaplan-Meier estimator.

For each threshold probability (*p*_*t*_), net benefit (NB) was computed as:

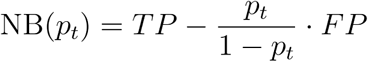

The threshold probability represents the minimum recurrence risk at which treatment would be recommended and the strategy with the highest net benefit is considered optimal.

### Statistical analysis

Statistical analyses were performed using Python 3.10 and R 4.0+ via rpy2.^28^ Data management and manipulation were conducted using pandas (v1.5)^29^ and numpy.^30^

### Survival analysis

Risk stratification and prognostic accuracy were measured using Harrell’s concordance index. Risk score was analyzed both as a continuous variable calculating hazard ratio per 0.1 unit increase in score as well as a dichotomized variable using a 10% threshold. A 10% threshold was chosen, yielding a partition of roughly two-thirds of patients as low-risk and one-third as high-risk. Continuous scores were used for multivariate Cox models; dichotomized scores were used for univariate Cox models.

Kaplan-Meier curves^31^ were estimated using the KaplanMeierFitter class from lifelines,^32^ with 95% confidence intervals estimated using Greenwood’s formula. Statistical significance was assessed using dataset-stratified univariate Cox proportional hazards models.^33^ To assess the impact of multiple inputs on the outcome, multivariate Cox proportional hazard models were fit. Multivariate models included tumor stage, nodal stage, and grade. Multivariate Cox models were stratified by dataset. Cox proportional hazards regression models were fitted using CoxPHFitter from lifelines and overall model significance was assessed using likelihood ratio tests. In multivariable Cox models, rare categorical levels (<10 observations) were aggregated into a single ‘other’ category to avoid sparse-level separation and ensure numerical stability. Hazard ratios for T stage categories were subsequently re-expressed relative to T1 via linear contrasts. Median follow up time was calculated using the reverse Kaplan-Meier method.

### Model calibration

Time-dependent model calibration of predicted 5-year recurrence risk probabilities was assessed by comparing Kaplan-Meier estimated recurrence probabilities within quantiles of predicted risk. Calibration curves were generated at 5 years and the agreement between predicted and observed risk was measured using the brier_score function from scikit-survival.^34^

### Meta-analysis

Hazard ratios were pooled using random effect models and bootstrapped using 1000 iterations.^35^ The meta-analysis was performed using the R meta package (v8.2-1)^36^ accessed via rpy2. Concordance index was computed with 1000 bootstrap iterations to generate confidence intervals.

### Data visualization

All figures were generated using matplotlib.^37^

## Supplemental Figures

**Supplemental Figure 1:**
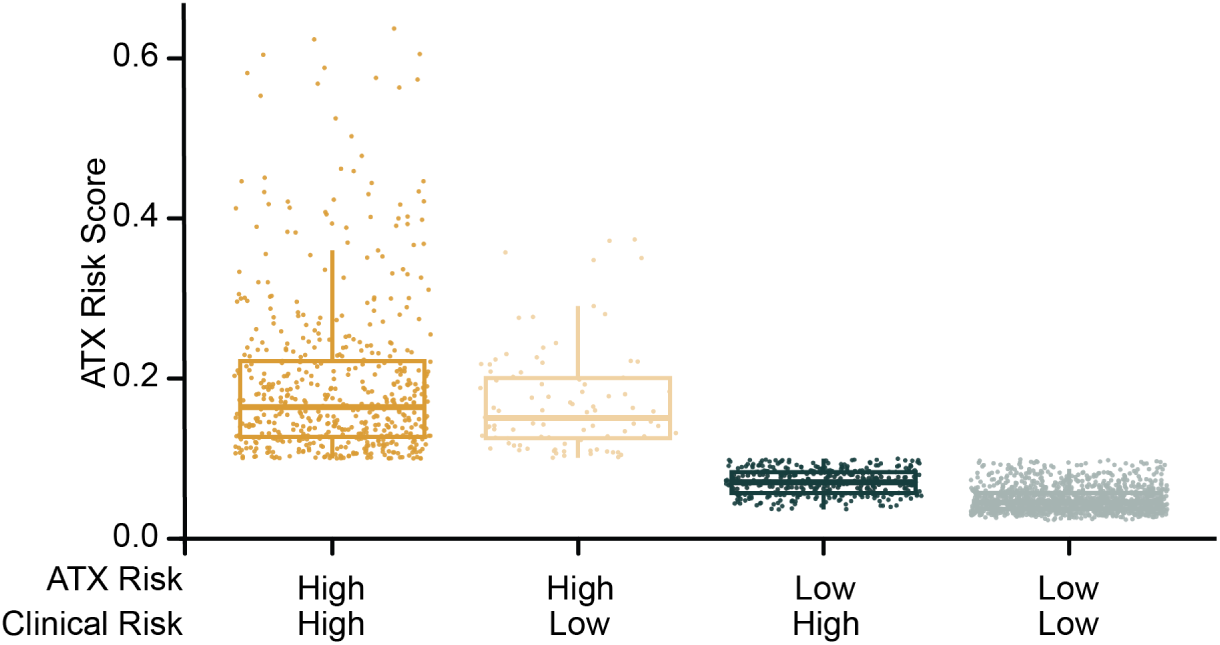
Distribution of ATX RISK scores stratified by ATX and clinical risk categories. Each point represents an individual patient. ATX scores are heterogeneous across clinical risk categories.

**Supplemental Figure 2:**
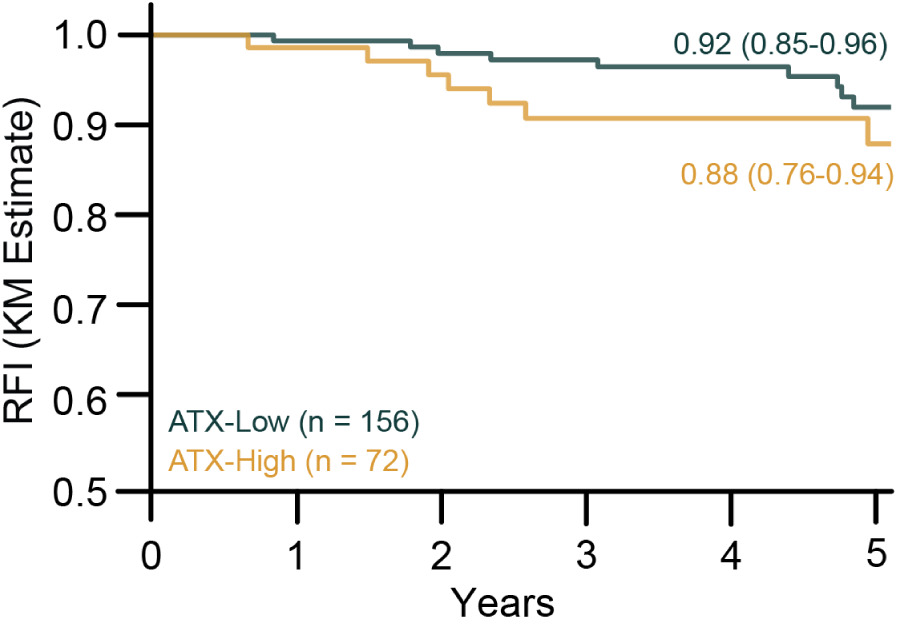
Kaplan-Meier plot of recurrence-free interval in T2N0 patients, stratified by ATX risk score (cut-off = 0.1).

## Data Availability

Data from The Cancer Genome Atlas (TCGA) are available via The Cancer Imaging Archive (TCIA) under DOI: 10.7937/K9/TCIA.2016.AB2NAZRP. The Providence dataset is available through Nightingale under DOI: 10.48815/N5159B. All other datasets analyzed in this study are private or proprietary and are not publicly available.

## Code Availability

The model can be accessed for non-commercial research upon request. A jupyter notebook for data analyses is also available upon request.

## Acknowledgements

This study makes use of data generated by the Molecular Taxonomy of Breast Cancer International Consortium. Funding for the project was provided by Cancer Research UK and the British Columbia Cancer Agency Branch. We also acknowledge the primary METABRIC publication.^38^ Tissues and samples were received from the Australian Breast Cancer Tissue Bank which is generously supported by the National Health and Medical Research Council of Australia, The Cancer Institute NSW and the National Breast Cancer Foundation. The tissues and samples are made available to researchers on a non-exclusive basis. The authors wish to acknowledge the roles of the Barts Cancer Now Tissue Bank in collecting and making available the samples and/or data, and the patients who have generously donated their tissues and shared their data to be used in the generation of this publication. Biosamples were obtained from the Wales Cancer Bank (DOI: http://doi.org/10.5334/ojb.46) which is funded by Health and Care Research Wales. Other investigators may have received specimens from the same subjects. We acknowledge the contributions (contributed materials) of the Northern Ireland Biobank in all academic outputs incorporating, or resulting from the use of, the material. Biological materials were provided by the Ontario Tumour Bank, which is supported by the Ontario Institute for Cancer Research through funding provided by the Government of Ontario. The views expressed in this publication are the views of the authors and do not necessarily reflect those of the Government of Ontario. The results published here are in part based upon data generated by the TCGA Research Network: http://cancergenome.nih.gov/. We acknowledge the contribution of the University of Chicago as a source of data used in this study.

## Author Contributions

AB, NPM, KJG, and JW conceived the study. NPM, CT, JC, KZ, KJG, JW assisted with data collection and analytical methodology development. AB, NPM, CT, KJG, and JW designed and performed the experiments. AB, NPM, CT, KJG, and JW wrote the paper. NPM, CT, CM, AAD, EDC, AB, JMA, JC, KZ, KJG, and JW critically reviewed the manuscript and provided final approval.

